# Analytical performance of a highly sensitive system to detect gene variants using next-generation sequencing for lung cancer companion diagnostics

**DOI:** 10.1101/2021.10.13.21264976

**Authors:** Kikuya Kato, Jiro Okami, Harumi Nakamura, Keiichiro Honma, Yoshiharu Sato, Seiji Nakamura, Yoji Kukita, Shin-ichi Nakatsuka, Masahiko Higashiyama

## Abstract

The recent increase in the number of molecular-targeted agents for non-small cell lung carcinoma and the corresponding target genes has led to the demand for the simultaneous testing of multiple genes. Although gene panels that use next-generation sequencing (NGS) are ideal for this purpose, conventional panels require a high tumor content, and biopsy samples often do not meet this requirement. We developed a new NGS panel, called a compact panel, to accommodate biopsy samples without restricting tumor content. The panel was characterized by high sensitivity, with detection limits for mutations of 0.14%, 0.20%, 0.48%, 0.24%, and 0.20% for *EGFR* exon 19 deletion, L858R, T790M, *BRAF* V600E, and *KRAS* G12C, respectively. Mutation detection also had a high quantitative ability, with correlation coefficients ranging from 0.966 to 0.992. The threshold for fusion detection was 1%. The panel exhibited good concordance with the approved tests. Identity rates are as follows: *EGFR* positive, 100% (95% confidence interval, 95.5-100); *EGFR* negative, 90.9 (82.2-96.3); *BRAF* positive, 100 (59.0-100); *BRAF* negative, 100 (94.9-100); *KRAS* G12C positive, 100 (92.7-100); *KRAS* G12C negative, 100 (93.0-100); *ALK* positive, 96.7 (83.8-99.9); *ALK* negative, 98.4 (97.2-99.2); *ROS1* positive, 100 (66.4-100); *ROS1* negative, 99.0 (94.6-100); *MET* positive, 98.0 (89.0-99.9); *MET* negative 100 (92.8-100); *RET* positive, 93.8 (69.8-100); *RET* negative, 99.8 (94.9-100). The analytical performance showed that the compact panel could handle various types of biopsy samples obtained by routine clinical practice without requiring strict pathological monitoring, as in the case of conventional NGS panels.

## Introduction

One of the major paradigms in modern cancer therapy is the prediction of the efficacy of anticancer agents based on genomic information from the patient. The foremost example of this paradigm is therapy for advanced non-small-cell lung carcinoma (NSCLC). Molecular-targeted agents are selected for patients based on mutations in target genes, a process called companion diagnostics. The era of precision medicine began with the discovery of a correlation between *EGFR* mutations and efficacy ^1^. Since its discovery, the number of molecular-targeted agents for NSCLC has continuously increased. Current molecular-targeted agents with companion diagnostics include osimertinib ^2^, alectinib ^3^, crizotinib ^4^, dabrafenib plus trametinib ^5^, tepotinib ^6^, selpercatinib ^7^, and sotorasib ^8^ for EGFR, ALK, ROS1, BRAF, MET, RET, and KRAS, respectively. Other agents targeting EGFR with exon 20 insertion ^9^ will be available in the near future.

Previously, only EGFR tyrosine kinase inhibitors were available and diagnostic tests for one gene were sufficient for practical use. However, with an increasing number of agents, there is now a strong demand for testing multiple genes simultaneously. In this context, gene panels, next-generation sequencing (NGS) panels, namely analytical/diagnostic systems to detect variants in multiple cancer-related genes using NGS, such as Foundation One CDx (Foundation Medicine, Inc.) and Oncomine Dx Target Test (Thermo Fisher Scientific), should be useful.

However, these panels are not necessarily optimized for diagnostic purposes. The main limitation is the restriction of tumor samples; high tumor content (>20%) is required for panel tests. This restriction prevents a considerable proportion of the samples from being tested, and such samples must be subjected to single-gene tests based on real-time polymerase chain reaction (PCR). These NGS panels were originally developed for samples collected for research purposes and are not optimized for testing various types of clinical samples in the real world. This shortcoming is mainly due to the low sensitivity of variant detection. We developed an NGS panel, named the compact panel, for companion diagnostics of NSCLC. The compact panel achieved a higher sensitivity than conventional NGS panels and single-gene tests. This panel was approved as a medical device by the Ministry of Health, Labour, and Welfare of Japan on November 16, 2022.

## Results

### Design of the compact panel

The compact panel limits the number of target genes to improve the flexibility of the panel design. The panel consisted of four modules: two DNA modules and two RNA modules, using DNA and RNA as templates, respectively. The target mutations were as follows: DNA module I for *EGFR* exon 19 deletion, *EGFR* L858R, *BRAF* V600E, and *KRAS* G12C; DNA module II for other *EGFR* mutations, *Her2* exon 20, and *MET* exon 14 skipping/amplification; RNA module I for *ALK* and *MET* fusion genes; and RNA module II for *ROS1* and *RET* fusion genes. The design of the compact panel is presented in Table 1. Detailed lists of *ALK, ROS1*, and *RET* fusion variants are provided in Supplementary File 1. The modular structure of the panel simplifies the process of obtaining official authorization because the analytical performance can be evaluated for each module without the need to reevaluate the entire panel. Additionally, the replacement or addition of modules can help address new diagnostic genes in the future.

**Table 1.**
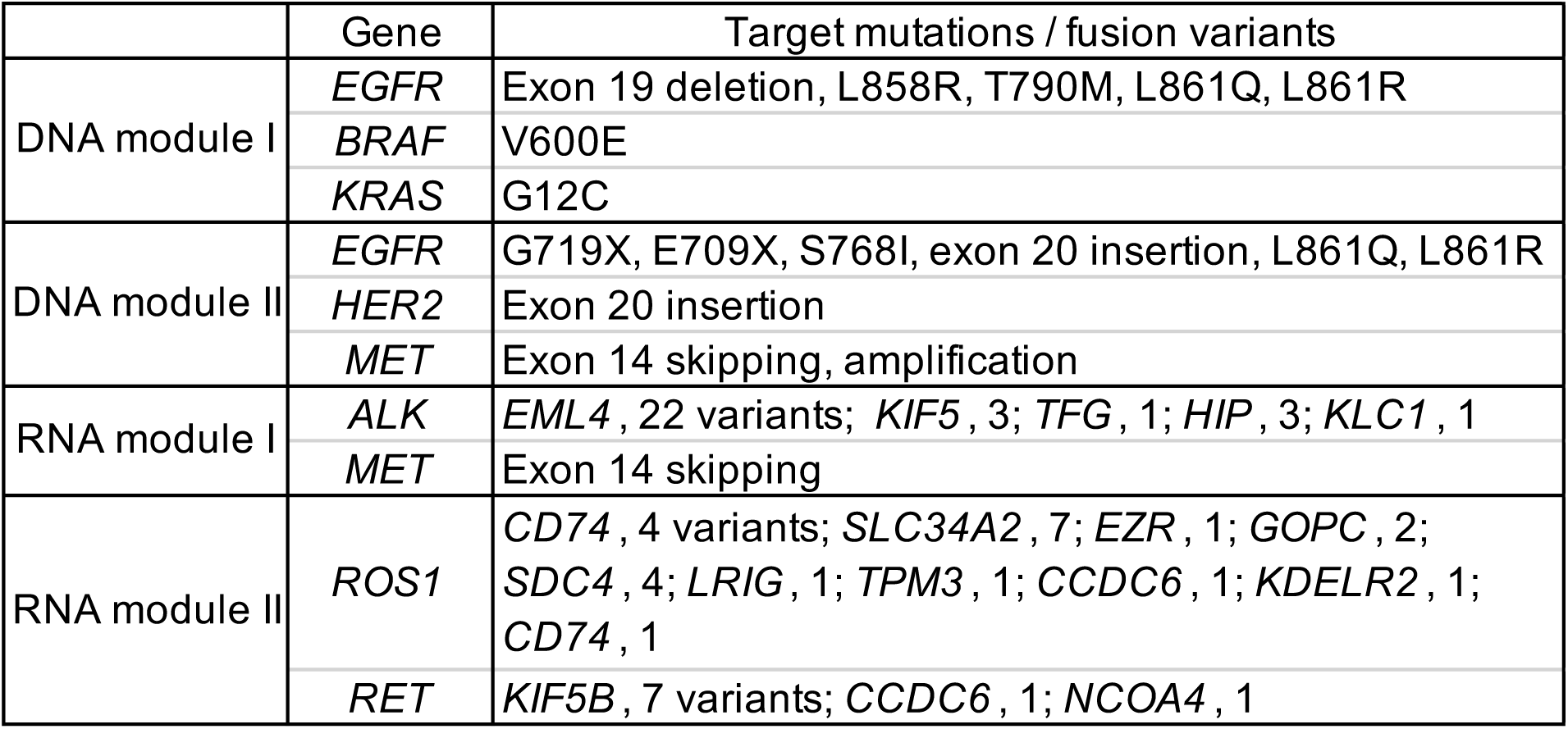
Modules of the compact panel. RNA module I includes *HGPRT1* for the control of amplification.

To increase the sensitivity, we applied different strategies for mutation and fusion. To detect mutations, templates were amplified from genomic DNA, and deep sequencing was performed, that is, repeated sequencing of target regions. Because the sensitivity depends on the number of sequence reads ^10^, we set the number to 5,000 instead of 700 with the Oncomine Dx Target Test. To detect fusions, multiplex PCR was used to amplify junctions of known variants, using RNAs as a template. Multiplex PCR is designed to optimize the amplification of fusions, thus reducing the number of normal genes among the amplified products. Consequently, a high sensitivity was achieved.

PCR primers were designed so that the sizes of the amplified products were <100 base pairs except two amplicons, which allowed PCR amplification using deteriorated DNA/RNA templates. All primer sequences are listed in Supplementary File 2. Derivatives of KOD DNA polymerase, an archaeal family-B DNA polymerase, were used for PCR amplification. Because the 3’–5’ proofreading exonuclease of archaeal family-B DNA polymerase hinders the copying of template strand deaminated bases ^11^, false positive mutations generated during formaldehyde fixation are expected to be excluded.

### Sensitivity of mutation detection using DNA as a template

The thresholds for mutation detection were set using an anomaly detection algorithm ^10^. For anomaly detection, the probability of false positives was estimated from the measured values of normal DNA, assuming a Poisson distribution. The threshold values were set so that the probability of false positives was 10^−10^. These values were defined by allele frequency (Table 2).

**Table 2.** Sensitivity test of the compact panel. The number of samples analyzed was 24, except *ALK* false negative (48 samples)

|  | Threshold of detection<br>DNA, % allele frequency;<br>RNA, TM score | Sensitivity test |  |
| --- | --- | --- | --- |
|  |  | False negative | False positive |
| <u>DNA module</u> |  |  |  |
| *EGFR* exon 19 del | 0.14 | 0 | 0 |
| *EGFR* L858R | 0.20 | 0 | 0 |
| *EGFR* T790M | 0.48 | 0 | 0 |
| *BRAF* V600E | 0.24 | 0 | 0 |
| *KRAS* G12C | 0.20 | 0 | 0 |
| <u>RNA module</u> |  |  |  |
| *ALK* fusion | 188 | 0 | 1 |
| *ROS1* fusion | 32 | 0 | 1 |
| *RET* fusion | 18 | 0 | 0 |
| *MET* exon 14 skipping | 28 | 0 | 0 |

To assess the sensitivity of DNA module I, we measured 24 samples with 1% and 0% mutant allele DNA for each test. There were no false positives, i.e., mutation positives in 0% mutated DNA, and no false negatives, i.e., mutation negatives in 1% mutated DNA (Table 2).

### Sensitivity of fusion detection using RNA as a template

The thresholds for fusion detection were set using an anomaly detection algorithm. The threshold values were set so that the probability based on the Poisson distribution of false positives was 10^−5^. The values were defined by the TM (tumor mutation) score, namely the number of positive reads per 100,000 reads, and are shown in Table 2.

Fusion-positive samples for sensitivity tests were prepared so that 1% of the RNA was derived from fusion-positive cells. In the sensitivity tests, we measured 24 or 48 fusion-positive and fusion-negative RNA samples for each test. The false positive/negative rates were <0.5% (Table 2).

### Quantification of mutation detection

Deep sequencing enables excellent quantification of mutated alleles. The quantitative ability of DNA module I was examined using artificial DNA samples prepared such that 1−8% of the total DNA consisted of mutant alleles. The results are shown in Figure 1.

**Figure 1.**
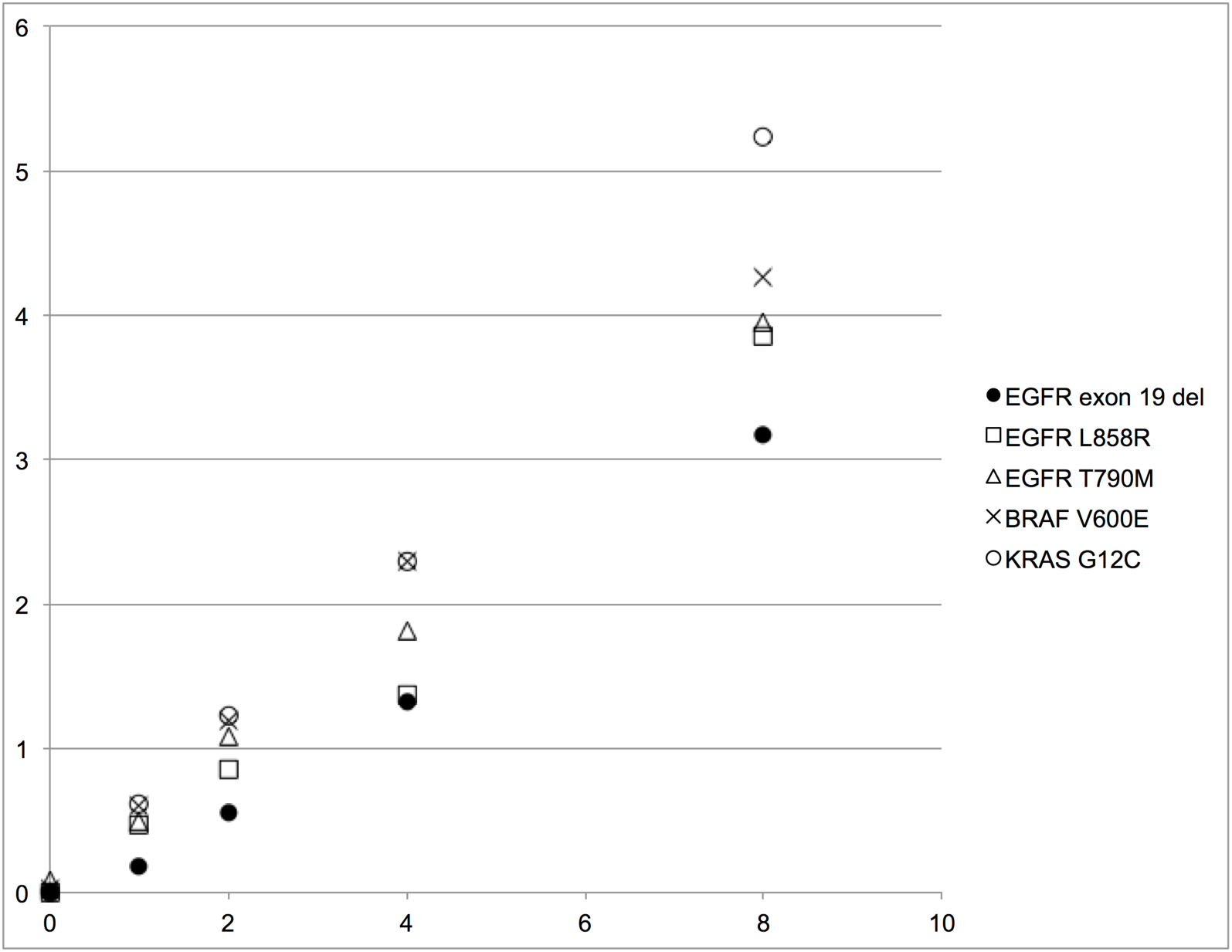
The quantitative ability of DNA module I. Horizontal axis, mutation allele frequency of artificial DNA (%); vertical axis, mutation allele frequency measured with the compact panel (%). Each data point is an average of eight samples.

There was a very good linearity between the inoculated numbers of mutant alleles and the observed mutant allele frequency deduced from deep sequencing. The correlation coefficients were 0973, 0.972, 0.966, 0.992, and 0.991 for *EGFR* exon 19 deletion, *EGFR* L858R, *EGFR* T790M, *BRAF* V600E, *and KRAS* G12C, respectively.

### Concordance with conventional diagnostic tests

The performance of the compact panel was compared with that of approved diagnostic tests. The reference diagnostic tests were as follows: *EGFR*, Cobas EGFR Mutation Test v2 (Roche Diagnostics K.K.); *ALK*, Histofine ALK iAEP kit (Nichirei Bioscience Inc.) and Vysis ALK Break Apart FISH probe kit (Abbott Laboratories); *MET*, ArcherMET (Archer DX, Inc.); *KRAS* G12C, therascreen KRAS RGQ PCR Kit; *BRAF* and *RET*, Oncomine Dx Target Test. Formalin-fixed paraffin-embedded (FFPE) NSCLC samples were simultaneously analyzed using a compact panel and a reference test, and the concordance of both tests was examined. The results are presented in Table 3. This good concordance proved the feasibility of the compact panel for practical use.

**Table 3.** Concordance of the compact panel with approved diagnostic tests. Parentheses in identity rates indicate a 95% confidence interval.

|  |  | Compact panel positive | Compact panel negative | Positive identity rate | Negative identity rate | Reference test |
| --- | --- | --- | --- | --- | --- | --- |
| *EGFR* | Reference test positive | 73 | 0 | 100<br>(95.5-100) | - | Cobas EGFR Mutation Test v2 |
|  | Reference test negative | 7 | 70 | - | 90.9<br>(82.2-96.3) |  |
| *BRAF*<br>V600E | Reference test positive | 7 | 0 | 100<br>(59.0-100) | - | Oncomine Dx Target Test |
|  | Reference test negative | 0 | 70 | - | 100<br>(94.9-100) |  |
| *KRAS*<br>G12C | Reference test positive | 49 | 0 | 100<br>(92.7-100) | - | therascreen KRAS RGQ PCR Kit |
|  | Reference test negative | 0 | 51 | - | 100<br>(93.0-100) |  |
| *ALK* | Reference test positive | 29 | 1 | 96.7<br>(83.8-99.9) | - | Histofine ALK iAEP kit / Vysis ALK Break Apart FISH probe kit |
|  | Reference test negative | 11 | 692 | - | 98.4<br>(97.2-99.2) |  |
| *ROS1* | Reference test positive | 9 | 0 | 100<br>(66.4-100) | - | OncoGuide AmoyDx ROS1 |
|  | Reference test negative | 1 | 99 | - | 99.0<br>(94.6-100) |  |
| *MET* | Reference test positive | 48 | 1 | 98.0<br>(89.0-99.9) | - | ArcherMET |
|  | Reference test negative | 0 | 50 | - | 100<br>(92.8-100) |  |
| *RET* | Reference test positive | 15 | 1 | 93.8<br>(69.8-100) | - | Oncomine Dx Target Test |
|  | Reference test negative | 0 | 70 | - | 99.8<br>(94.9-100) |  |

Seven discordant *EGFR* samples (compact panel positive and reference test negative) were analyzed using digital PCR, and all samples were found to be mutation-positive, consistent with the results of the compact panel. One discordant *ALK* sample was analyzed with the non-overlapping integrated read sequencing system (NOIR-SS) ^12^ and it was found to be an *ALK* fusion with *C2ORF71*, a rare fusion type not covered in both the compact panel and Oncomine Dx Target Test.

### Incidence of mutations and fusions

NSCLC samples stored at the Osaka International Cancer Institute were screened using a compact panel and the incidence of mutations and fusions at the population level was determined (Table 4). The figures were compared to those obtained from the Oncomine Comprehensive Assay in a nationwide project named Scrum-Japan ^13^. *EGFR* mutation-positive samples were excluded from the Osaka International Cancer Institute cohort because Scrum-Japan collected only *EGFR* mutation-negative samples, and the observed incidence was higher with *KRAS, BRAF, ALK*, and *MET* in the Osaka International Cancer Institute cohort and with *ROS1* in the Scrum-Japan cohort.

**Table 4.** Frequencies of mutations and fusions among *EGFR*-mutation negative samples. Parentheses in percent frequency indicate a 95% confidence interval. Samples from the Osaka International Cancer Institute are obtained from October 26, 2015, to August 24, 2020. The numbers of *BRAF* V600E and *KRAS* G12C samples are 11 and 77, respectively.

|  |  |  |  |  |
| --- | --- | --- | --- | --- |
|  | Osaka International Cancer Institute |  | Scrum-Japan |  |
| Assay system | Compact panel |  | Oncomine comprehensive assay |  |
| Total number of samples | 827 |  | 3919 |  |
| Gene | Number of variant-positive samples | Percent frequency | Number of variant-positive samples | Percent frequency |
| *KRAS* | 230 | 27.8 (24.8-31.0) | 382 | 9.7 (8.8-10.7) |
| *BRAF* | 55 | 6.7 (5.1-8.6) | 97 | 2.5 (2.0-3.0) |
| *ALK* | 41 | 5 (3.6-6.7) | 97 | 2.5 (2.0-3.0) |
| *ROS1* | 10 | 1.2 (0.6-2.2) | 142 | 3.6 (3.1-4.3) |
| *MET* | 56 | 6.8 (5.2-8.7) | 98 | 2.5 (2.0-3.0) |
| *RET* | 18 | 2.2 (1.3-3.4) | 100 | 2.6 (2.1-3.1) |

## Discussion

Companion diagnostics has long been performed using real-time PCR-based tests for individual genes. However, due to the increasing number of target genes, there is a strong demand for the simultaneous testing of multiple genes. In this context, the simultaneous analysis of multiple genes using NGS is ideal. In the USA, NGS panels were performed in 44% of patients with NSCLC who require genetic test ^14^, and are becoming indispensable for NSCLC treatment.

However, the introduction of NGS panels into Japanese medical practice is not straightforward. In the USA, NSCLC samples subjected to genetic tests are usually obtained by core needle biopsy or transthoracic fine needle aspiration biopsy guided by computed tomography or ultrasound ^15^. In contrast, biopsy using bronchoscopy is more popular in Japan. This difference in biopsy practice causes a unique problem that is not apparent in the medical environment in the USA. In Japan, the feasibility of NGS panels is generally lower than that of other diagnostics in clinical practice, especially regarding nonsurgical biopsy ^16^. The success rate of the Oncomine Dx Target Test is influenced by tissue size and tumor cell count ^17, 18^. Therefore, strict pathological monitoring is necessary to select the samples subjected to the test; consequently, a considerable fraction of the samples were excluded from the test.

This issue can be addressed by introducing a new NGS panel with high sensitivity. We developed a compact panel, an NGS panel that accommodates various types of NSCLC samples without requiring strict pathological monitoring. The sensitivity of the compact panel suggested the detection of mutation/fusion in samples with 1% tumor content. The confirmed detection limit of DNA module I of the compact panel, which is 1% mutation allele frequency, is lower than that of the Oncomine Dx Target Test, which is 6−13%, and Cobas EGFR Mutation Test v2, which is 1.26−6.81% (FDA Summary of Safety and Effectiveness Data). Because the amount of DNA/RNA required is only 5 ng, a smaller amount of tissue would need to be obtained by routine biopsy practice. Although the compact panel was originally designed for the Japanese medical environment, such high-sensitivity NGS panels would also benefit other medical environments.

The compact panel could handle various sample types, including cytological samples. The preliminary application of the compact panel showed stable performance on cytological samples^19-21^, which was not possible with conventional NGS panels.

Formalin fixation, which may deteriorate nucleic acid quality, is not necessary for genetic testing. The ammonium sulfate solution allows the efficient preservation of RNA/DNA for several days at room temperature ^22^ and is expected to replace formalin fixation. This would be particularly useful as a washing solution for samples obtained by cytodiagnostic brushing or curette washing.

## Materials and methods

### DNA and RNA samples

#### Sensitivity tests

DNA samples, including various fractions of mutant alleles, were prepared using the following solid tumor analysis reference standards (Horizon Discovery Ltd.): HDx FFPE EGFR e19 del. 50%; HDx FFPE EGFR L858R 50%; FFPE EGFR T790M 50%; HDx FFPE BRAF V600E 50%. To adjust allele frequencies, wild-type FFPE genomic DNA from the lungs, including bronchiole (Cureline Inc.), was used. RNA samples, including fusions, were constructed from *ALK-RET-ROS1*-targeted FFPE RNA Fusion Reference Standards (Horizon Discovery Ltd.). For *MET* exon 14 skipping, synthesized DNA was used, skipping the need for reverse transcription. Wild-type FFPE RNA from the lungs, including bronchioles (Cureline, Inc.), was used to adjust the RNA concentration.

#### Concordance tests

FFPE NSCLC samples were obtained by surgical resection and stored at the Osaka International Cancer Institute. DNA and RNA were purified from the samples using a Maxwell RSC instrument (Promega Corporation). Studies using clinical samples were approved by the ethics committees of the Osaka International Cancer Institute and Nara Institute of Science and Technology.

### Library preparation and sequencing of DNA modules

PCR amplification was performed in 50 μL of the reaction mixture containing 1 × buffer (Toyobo, Inc.), 0.2 mM dNTPs, 1.5 mM Mg_2_SO_4_, 5 ng of DNA purified from FFPE NSCLC, 0.3 μM each of the primer mixture and 0.02 U of KOD -Plus-Neo (Toyobo, Inc.). Forty cycles were performed at 98 °C for 10 s and 62 °C for 30 s. After purification with AMPure XP (Beckman Coulter Life Sciences), the amplified products were subjected to library construction using the MiSeq System (Illumina, Inc.) and the GenNext NGS Library Prep Kit (TOYOBO, Inc.). To discriminate individual samples, index sequences were introduced using the TruSeq DNA Single Indexes Set A or TruSeq DNA CD Indexes Set (Illumina, Inc.). Sequencing was performed using a MiSeq System (Illumina, Inc.). The minimum number of reads per fragment was 5,000.

### Library preparation and sequencing of RNA modules

Reverse transcription was performed in 20 μL of a reaction mixture containing 1 × ReverTra buffer (Toyobo, Inc.), 1 mM dNTPs, 10 ng of RNA purified from FFPE NSCLC, and 100 U of ReverTra Ace (Toyobo, Inc.). After denaturation with RNA and a 9-base random primer (Toyobo, Inc.) at 65 °C for 5 min, the reaction mixture was incubated at 30 °C for 10 min and then at 42 °C for 60 min. PCR amplification was performed in 50 μL of reaction mixture containing 1 × buffer (Toyobo, Inc.), 0.4 mM dNTPs, 1.5 mM Mg_2_SO_4_, the 20 μl reaction mixture mentioned above, 0.25 μM each of the primer mixtures, and 1 U of KOD -Fx-Neo (Toyobo, Inc.). Forty cycles were performed at 98 °C for 15 s, 60 °C for 30 s, and 68 °C for 10 s followed by an extension at 68 °C for 1 min. After purification with AMPure XP (Beckman Coulter Life Sciences), the amplified products were subjected to library construction and sequencing, as described in the previous section. The minimum number of reads required per sample was 300.

### Analysis of discordant samples

Digital PCR was performed using the QX200 Droplet Digital PCR System (Bio-Rad Laboratories, Inc.) following the supplier’s protocol. The NOIR-SS assay was performed as previously described ^12^.

## Data Availability

All data produced in the present study are available upon reasonable request to the authors

## Supporting information

Supplementary File 1. List of ALK, ROS1, and RET fusion variants.

Supplementary File 2. Sequences of primers for sequencing template preparation.

## Acknowledgments

The authors thank Miho Ishii, Yuki Mori, Yui Nose, Yumi Ueda, and Kazuya Taniguchi for their laboratory work. The authors also thank Hiroyuki Sato and Motohiko Tanino for data analysis. The authors are profoundly grateful to Yuji Horikawa for the administration.

